# Comparative Analysis of Multimodal Large Language Models GPT-4o and o1 vs Clinicians in Clinical Case Challenge Questions

**DOI:** 10.1101/2025.06.22.25330068

**Authors:** Jaewon Jung, Hyunjae Kim, SungA Bae, Jin Young Park

**Affiliations:** Department of Medicine, Yonsei University College of Medicine, Seoul, Republic of Korea; Department of Biomedical Systems Informatics, Yonsei University College of Medicine, Yongin, Republic of Korea; Division of Cardiology, Department of Internal Medicine, Yonsei University College of Medicine, Yongin Severance Hospital, Yongin, Republic of Korea; Department of Psychiatry, Yongin Severance Hospital, Yonsei University College of Medicine, Yongin, Republic of Korea

**Author notes:** These authors contributed equally (Jaewon Jung, Hyunjae Kim).

**Keywords:** Multimodal large language model, GPT-4 Omni, o1, clinical decision-making, diagnostic accuracy, artificial intelligence

## Abstract

**Background:** Generative Pre-trained Transformer 4 (GPT-4) has demonstrated strong performance in standardized medical examinations but has limitations in real-world clinical settings. The newly released multimodal GPT-4o model, which integrates text and image inputs to enhance diagnostic capabilities, and the multimodal o1 model, which incorporates advanced reasoning, may address these limitations.

**Objective:** This study aimed to compare the performance of GPT-4o and o1 against clinicians in real-world clinical case challenges.

**Methods:** This retrospective, cross-sectional study used Medscape case challenge questions from May 2011 to June 2024 (n = 1,426). Each case included text and images of patient history, physical examination findings, diagnostic test results, and imaging studies. Clinicians were required to choose one answer from among multiple options, with the most frequent response defined as the clinician’s decision. Data-based decisions were made using GPT models (3.5 Turbo, 4 Turbo, 4 Omni, and o1) to interpret the text and images, followed by a process to provide a formatted answer. We compared the performances of the clinicians and GPT models using Mixed-effects logistic regression analysis.

**Results:** Of the 1,426 questions, clinicians achieved an overall accuracy of 85.0%, whereas GPT-4o and o1 demonstrated higher accuracies of 88.4% and 94.3% (mean difference 3.4%; *P* = .005 and mean difference 9.3%; *P* < .001), respectively. In the multimodal performance analysis, which included cases involving images (n = 917), GPT-4o achieved an accuracy of 88.3%, and o1 achieved 93.9%, both significantly outperforming clinicians (mean difference 4.2%; *P* = .005 and mean difference 9.8%; *P* < .001). o1 showed the highest accuracy across all question categories, achieving 92.6% in diagnosis (mean difference 14.5%; *P* < .001), 97.0% in disease characteristics (mean difference 7.2%; *P* < .001), 92.6% in examination (mean difference 7.3%; *P* = .002), and 94.8% in treatment (mean difference 4.3%; *P* = .005), consistently outperforming clinicians. In terms of medical specialty, o1 achieved 93.6% accuracy in internal medicine (mean difference 10.3%; *P* < .001), 96.6% in major surgery (mean difference 9.2%; *P* = .030), 97.3% in psychiatry (mean difference 10.6%; *P* = .030), and 95.4% in minor specialties (mean difference 10.0%; *P* < .001), significantly surpassing clinicians. Across five trials, GPT-4o and o1 provided the correct answer 5/5 times in 86.2% and 90.7% of the cases, respectively.

**Conclusions:** The GPT-4o and o1 models achieved higher accuracy than clinicians in clinical case challenge questions, particularly in disease diagnosis. The GPT-4o and o1 could serve as valuable tools to assist healthcare professionals in clinical settings.

## INTRODUCTION

Rapid advancements in artificial intelligence (AI) have transformed various sectors, including healthcare, where the importance of digital health technologies continues to grow.^1–3^ Among these technologies, large language models (LLMs) have demonstrated considerable potential in healthcare, particularly in answering medical questions, assisting with diagnoses, and automating administrative tasks.^4^ Generative Pre-trained Transformer 4 (GPT-4; OpenAI, San Francisco, CA, USA) is one of the most advanced LLMs, consistently achieving high scores on the United States Medical Licensing Examination and often exceeding the passing threshold.^5–7^ Despite these accomplishments, existing evaluations have primarily focused on text-only formats that often exclude or replace visual elements with descriptions. This limitation reduces GPT-4’s applicability in image-dependent specialties, such as radiology.

To address the limitations of unimodal LLMs, the recently released GPT-4 Omni introduces multimodal capabilities, enabling it to process and integrate both textual and visual information.^8^ This enhancement allows for a more comprehensive analysis of clinical cases, particularly in fields such as diagnostic and interventional radiology and nuclear medicine, where early studies have demonstrated its potential.^9, 10^ Despite these advancements, existing research has largely focused on simplified exam questions, and no studies have reported significant performance improvements over text-only models in medical examinations.^11, 12^ This gap highlights the need to evaluate the performance of multimodal models in clinical case settings that reflect the complexity of diagnostic decision-making encountered in clinical practice.

In parallel with these multimodal developments, OpenAI recently introduced the o1 model, which incorporates advanced reasoning capabilities designed to enhance safety and robustness.^13^ By utilizing chain-of-thought reasoning, o1 has demonstrated improved adherence to safety protocols in complex scenarios, effectively reducing the risk of generating harmful or biased content. These enhanced reasoning capabilities are particularly important in healthcare, where complex problem-solving and data interpretation are critical.^14^ Several studies have shown that the o1-preview outperforms both GPT-4 and GPT-4o models in clinical settings, highlighting its superior reasoning abilities in medical decision-making processes.^15–17^ However, the performance of the o1 model in complex real-world clinical cases remains unexplored due to its recent release. Accordingly, this study aimed to evaluate the clinical potential of two advanced large language models, GPT-4 Omni and o1, in solving complex clinical case challenge questions that incorporate both textual and visual data and to validate their effectiveness as diagnostic support tools compared with clinicians.

## RESULTS

### Question characteristics

We evaluated the performance of four language models, GPT-3.5 Turbo, GPT-4 Turbo, GPT-4 Omni, and o1, using a dataset of 1,426 cases. Of these, 917 questions included images, resulting in 1,475 images. **Figure S1 in the Supplementary Appendix** shows the quiz samples. The number of answer options per question ranged from two to nine, with the majority of cases (97.2%) offering either four or five answer options: 906 cases had four options and 480 had five. The random chance of selecting the correct answer was 23.1%.

### Overall accuracy and model comparison

Clinicians demonstrated an accuracy of 85.0% across the entire dataset (**Figure 1A** and **Table S1 in the Supplementary Appendix**). In comparison, GPT-3.5 Turbo, GPT-4 Turbo, GPT-4o, and o1 achieved accuracies of 60.6% (95% confidence interval [CI], 58.1–63.1), 82.1% (95% CI, 80.1–84.0), 88.4% (95% CI, 86.8–90.1), and 94.3% (95% CI, 93.1–95.5), respectively. The performance of GPT-4o and o1 surpassed that of clinicians, demonstrating significantly higher accuracy (*P* = .005 and *P* < .001, respectively). In contrast, GPT-3.5 Turbo and GPT-4 Turbo performed significantly lower than the clinicians (*P* < .001 and *P* = .028, respectively).

**Figure 1.**
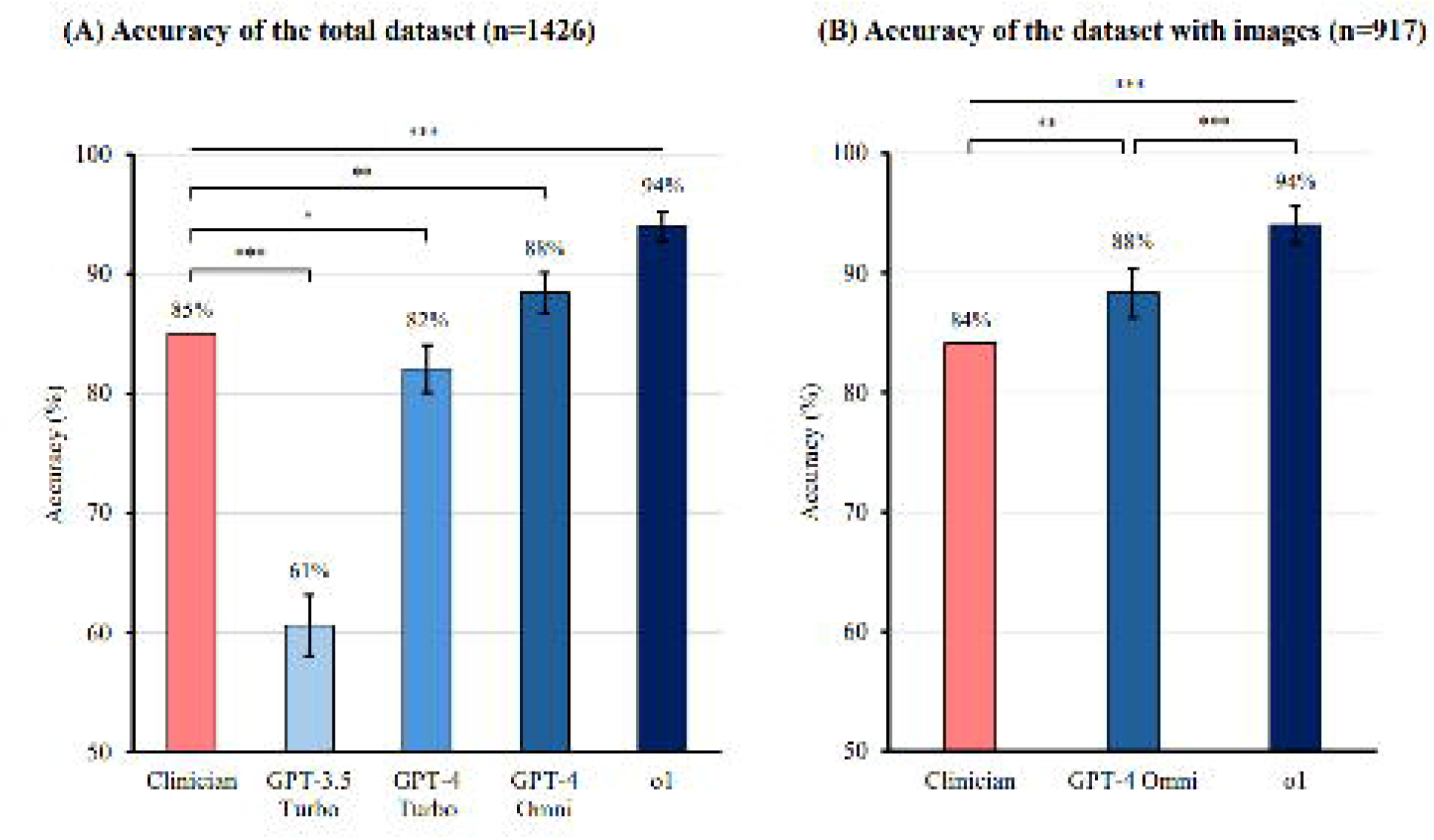
Overall performance of the GPT models. (A) Accuracy of the models across the entire dataset (n = 1,426). Clinicians achieved an accuracy of 85.0%, while GPT-3.5 Turbo, GPT-4 Turbo, GPT-4 Omni, and o1 achieved accuracies of 60.6%, 82.1%, 88.4%, and 94.3%, respectively. Statistical comparisons showed that GPT-4o and o1 significantly outperformed clinicians (\*\**P* < .01; \*\*\**P* < .001), while GPT-3.5 Turbo and GPT-4 Turbo performed significantly worse (\*\*\**P* < .001; \**P* < .05). Error bars represent 95% confidence intervals. (B) Accuracy of models for the dataset subset with images (n = 917). Clinicians achieved an accuracy of 84.1%, while GPT-4o achieved 88.3%, significantly surpassing clinicians (\*\**P* < .01). o1 demonstrated an accuracy of 93.9%, significantly outperforming both clinicians and GPT-4o (\*\*\**P* < .001 for both). Error bars represent 95% confidence intervals. GPT-4, Generative Pre-trained Transformer 4

To assess the potential impact of data exposure on model performance, we evaluated the accuracy of GPT-4o and o1 on datasets introduced both before (n = 1,219) and after (n = 207) the knowledge cutoff date of October 2023. Both models demonstrated comparable accuracy across the two time periods (*P* = .646 for GPT-4o and *P* = .249 for o1), indicating that exposure to the test data had minimal to no impact on model accuracies (**Figure S2 in the Supplementary Appendix**).

### Multimodal model performances on cases with images

In the subset of cases that included images (n = 917), clinicians achieved an accuracy of 84.1% (**Figure 1B**). GPT-4o performed better, with an accuracy of 88.3% (95% CI, 86.3–90.4), which was significantly higher than that of clinicians (*P* = .005). o1 exhibited the highest performance, achieving an accuracy of 93.9% (95% CI, 92.3–95.4), significantly surpassing both clinicians and GPT-4o (*P* < .001 for both).

### Performance by question category

The dataset was stratified based on the question category, with the diagnosis category comprising the largest number of questions (n = 530), followed by disease characteristics (n = 401), treatment (n = 305), and examination (n = 190) (**Table 1**). o1 demonstrated the highest accuracy across all question categories, consistently outperforming clinicians and other GPT models. Overall, it achieved 94.3% accuracy, significantly surpassing that of clinicians (85.0%, *P* < .001) and all other models.

**Table 1.**
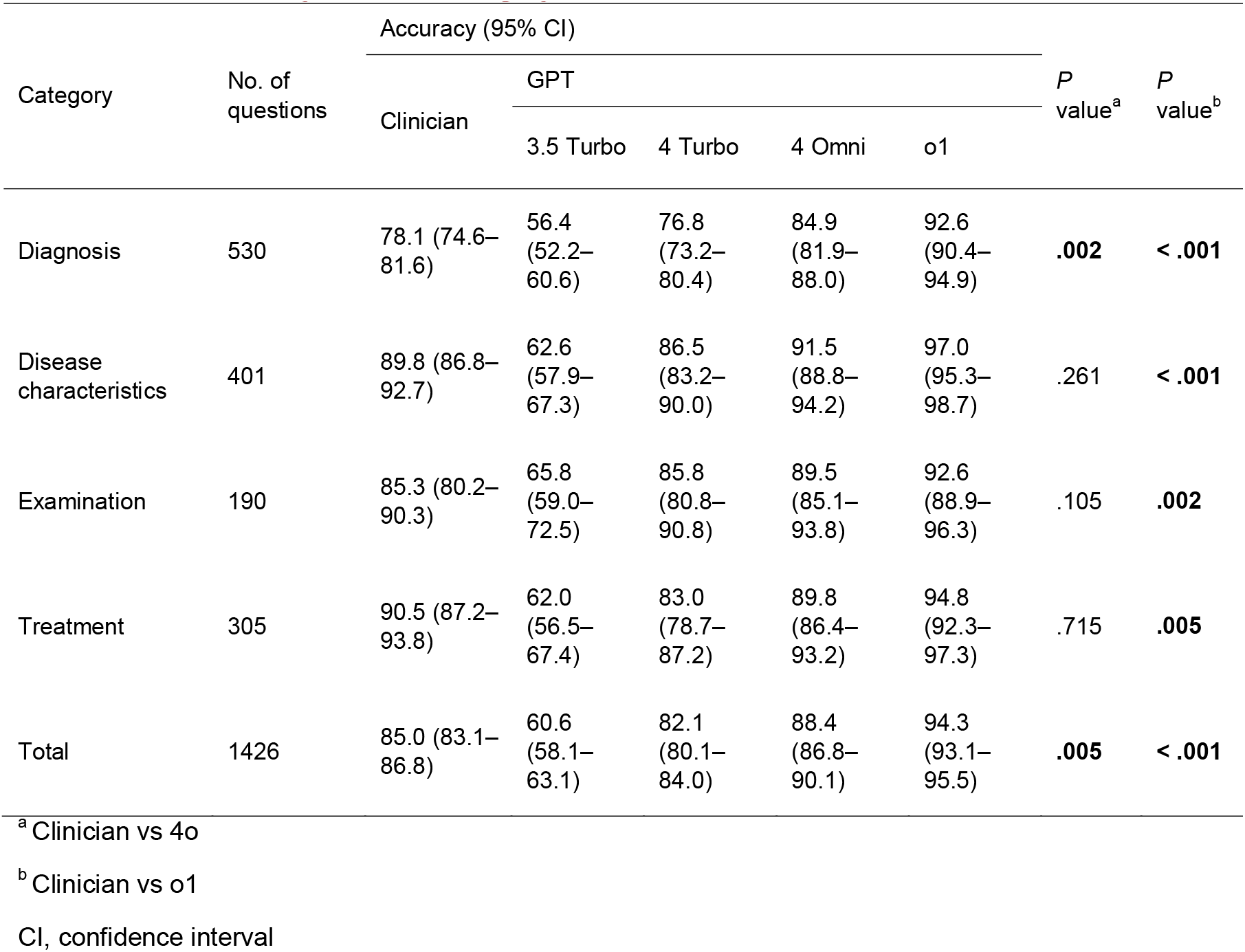
Performance by Question Category

In the diagnosis and disease characteristics categories, o1 achieved 92.6% and 97.0% accuracy, respectively, significantly outperforming clinicians (*P* < .001 for both). In the examination category, o1 achieved 92.6% accuracy, surpassing that of clinicians, who achieved 85.3% accuracy (*P* = .002). In the treatment category, o1 also outperformed clinicians, achieving 94.8% accuracy compared to 90.5% for clinicians (*P* = .005).

In the diagnosis category, GPT-4o significantly outperformed clinicians, achieving 84.9% accuracy compared to 78.1% for clinicians (*P* = .002). For categories related to disease characteristics and examination, GPT-4o also demonstrated higher accuracy than clinicians, although the differences were not statistically significant (*P* = .261 and *P* = .105, respectively). In the treatment category, clinicians showed greater accuracy than GPT-4o, but the difference was not statistically significant (*P* = .715).

### Performance by medical specialty

The dataset was stratified by medical specialty, with internal medicine comprising 766 questions, followed by minor specialties (n = 390), pediatrics (n = 108), major surgery (n = 87), and psychiatry (n = 75) (**Table 2**). o1 achieved 93.6% accuracy in internal medicine and 95.4% in minor specialties, which were statistically significant compared to clinicians (p < .001 for both). In major surgery, o1 outperformed clinicians with 96.6% accuracy, while clinicians achieved 87.4% accuracy (*P* = .030). In psychiatry, o1 achieved an accuracy of 97.3%, surpassing that of clinicians, who achieved 86.7% accuracy (*P* = .030). Clinicians demonstrated numerically higher performance in pediatrics than in o1, but the difference was not statistically significant (*P* = .798).

**Table 2.** Performance by Medical Specialty

| Category | No. of questions | Accuracy (95% CI) |  |  |  |  | *P* value<sup>a</sup> | *P* value<sup>b</sup> |
| --- | --- | --- | --- | --- | --- | --- | --- | --- |
|  |  | Clinician | GPT |  |  |  |  |  |
|  |  |  | 3.5 Turbo | 4 Turbo | 4 Omni | o1 |  |  |
| Internal medicine | 766 | 83.3<br>(80.6–85.9) | 59.0<br>(55.5–62.5) | 80.2<br>(77.3–83.0) | 86.9<br>(84.6–89.3) | 93.6<br>(91.9–95.3) | **.037** | **< .001** |
| Major Surgery | 87 | 87.4<br>(80.4–94.3) | 64.4<br>(54.3–74.4) | 79.3<br>(70.8–87.8) | 88.5<br>(81.8–95.2) | 96.6<br>(92.7–100.0) | .802 | **.030** |
| Pediatrics | 108 | 92.6<br>(87.7–97.5) | 61.1<br>(51.9–70.3) | 84.3<br>(77.4–91.1) | 90.7<br>(85.3–96.2) | 91.7<br>(86.5–96.9) | .617 | .798 |
| Psychiatry | 75 | 86.7<br>(79.0–<br>94.4) | 60.0<br>(48.9–<br>71.1) | 82.7<br>(74.1–<br>91.2) | 84.0<br>(75.7–<br>92.3) | 97.3<br>(93.7–<br>100.0) | .645 | **.030** |
| Minor | 390 | 85.4<br>(81.9–<br>88.9) | 62.8<br>(58.0–<br>67.6) | 85.6<br>(82.2–<br>89.1) | 91.5<br>(88.8–<br>94.3) | 95.4<br>(93.3–<br>97.5) | **.005** | **< .001** |
| Total | 1426 | 85.0<br>(83.1–<br>86.8) | 60.6<br>(58.1 –<br>63.1) | 82.1<br>(80.1–<br>84.0) | 88.4<br>(86.8–<br>90.1) | 94.3<br>(93.1–<br>95.5) | **.005** | **< .001** |

<sup>a</sup> Clinician vs 4o
<sup>b</sup> Clinician vs o1
CI, confidence interval

GPT-4o demonstrated superior accuracy in internal medicine, with 86.9% compared to 83.3% for clinicians (*P* = .037). Similarly, in minor specialties, GPT-4o outperformed clinicians, achieving 91.5% accuracy compared to 85.4% (*P* = .005). In addition, GPT-4o showed higher accuracy than clinicians in major surgeries, although the difference was not statistically significant (*P* = .802). In contrast, clinicians demonstrated higher accuracy in pediatrics and psychiatry, but these differences were not statistically significant (*P* = .617 and *P* = .645, respectively).

### Consistency of responses

GPT-4o and o1 were evaluated using the entire dataset over five independent trials to assess response consistency (**Figure 2**). GPT-4o achieved the correct answer 5/5 times in 86.2% of the cases, while o1 demonstrated a slightly higher consistency, with the correct answer 5/5 times in 90.7% of the cases. Notably, GPT-4o answered correctly at least once in 90.3% of the cases across the five trials, whereas o1 achieved at least one correct answer in 95.6% of the cases.

**Figure 2.**
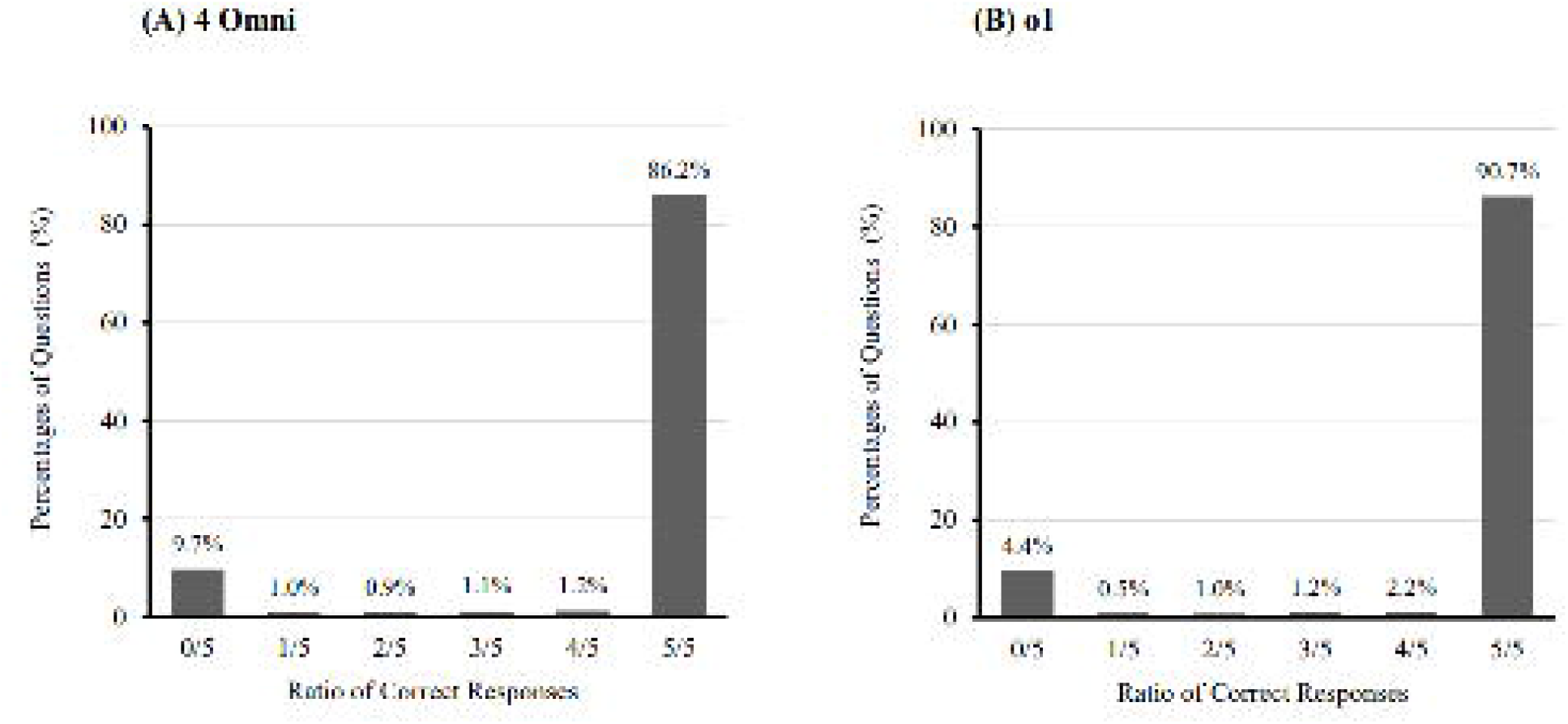
Response consistency of GPT-4o and o1. (A) Performance of GPT-4o across five independent trials, showing that 86.2% of cases had all correct responses (5/5). GPT-4o achieved at least one correct response in 90.3% of cases across the trials. (B) Performance of o1 across five independent trials, demonstrating slightly higher consistency, with 90.7% of cases achieving all correct responses (5/5). o1 achieved at least one correct response in 95.6% of cases across the trials. GPT-4, Generative Pre-trained Transformer 4

## DISCUSSION

### Principal results

This study demonstrated that recent GPT models, specifically o1 and GPT-4o, can outperform clinicians in clinical case challenges.

### Comparison with prior work

Previous studies using earlier models, such as GPT-3.5 Turbo and GPT-4 Turbo, generally found that the performance on medical cases was comparable to or below that of humans.^18, 19^ A prior study using Medscape clinical case challenges with a text-only model reported performance below 50%, highlighting significant limitations.^20^ In contrast, our study utilized multimodal models, resulting in substantial performance improvements. Both GPT-4o and o1 maintained high accuracy in cases involving images, demonstrating their ability to interpret various imaging types, including magnetic resonance imaging (MRI), computed tomography (CT), X-ray, ultrasound, and pathological images, across multiple specialties. Additionally, earlier research has shown that the o1-preview model outperformed the GPT-4 series in solving medical problems.^14, 16, 17^ Our findings further support this observation, as o1 outperformed GPT-4o in complex medical scenarios.

The accuracy of o1 remained above 90% across all question categories and medical specialties, with particular strength in diagnosis. The model also performed well in areas such as major surgery and psychiatry, with accuracy rates exceeding those of GPT-4o by more than 8%. Research suggests that o1’s high performance is driven by its “chain-of-thought” reasoning, which systematically breaks down complex problems, reduces hallucinations, and improves logical reasoning.^13, 14^ This capability is especially beneficial in clinical cases with abundant patient data, which often include extraneous information. In such contexts, o1’s ability to filter relevant insights and synthesize data across specialties aids in understanding disease progression and managing complex multisystem conditions.

The o1 model demonstrated stronger performance and greater consistency than GPT-4o in selecting the most accurate option from multiple choices. This finding contrasts with previous studies suggesting similar gains between the o1 and GPT-4 models in probabilistic reasoning and critical diagnosis identification.^21^ This divergence may be attributed to differences in prompt design and parameter tuning, which enabled o1 to achieve a significant improvement. However, it also suggests that o1’s performance may vary depending on the clinical setting.

### Integration into clinical workflows and application

While the study shows that AI models outperform clinicians in accuracy, their reasoning process remains a black box. Understanding how these models reach their conclusions and making this process interpretable for clinicians is a critical consideration for integration into clinical workflows. **Figure 3** illustrates how the AI model interprets imaging studies and synthesizes differential diagnoses. This approach enables clinicians to verify, understand, and critically assess AI recommendations, thereby facilitating safer and more informed decision-making.

**Figure 3.**
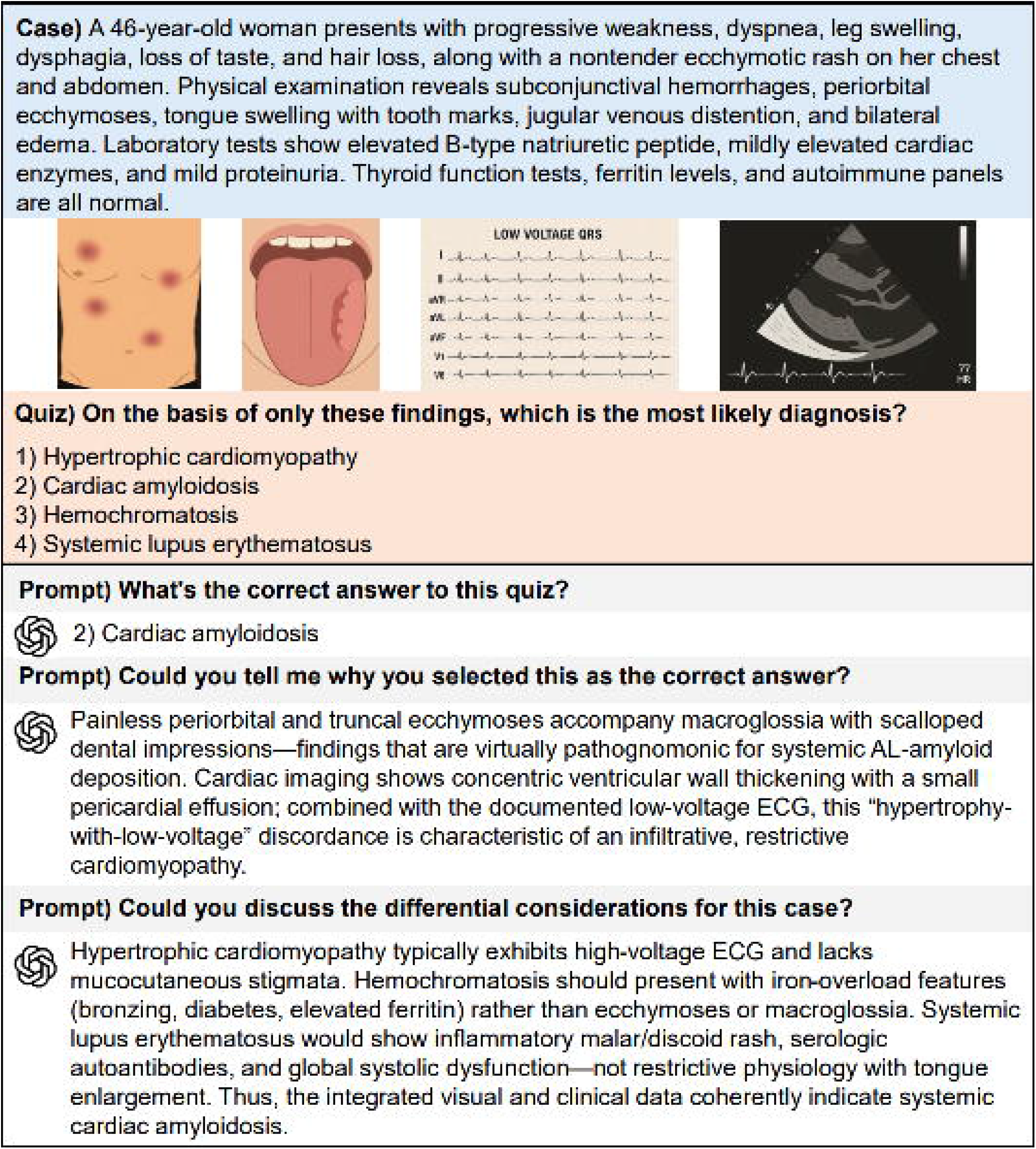
Multimodal Reasoning Process of GPT in a Clinical Case. This figure illustrates how the GPT model integrates textual and visual information to interpret a clinical case. The GPT model is prompted to identify the most likely diagnosis, explain its reasoning, and discuss differential diagnoses. GPT, Generative Pre-trained Transformer

In practical applications, GPT models may serve as valuable tools to support clinicians, particularly in settings where specialized expertise is limited. Clinicians are prone to diagnostic errors and may occasionally order unnecessary tests, influenced by factors such as financial incentives while lacking adequate tools to monitor such decisions.^22–25^ Recent models demonstrate potential in assisting with differential diagnosis and initial treatment planning, especially in fields outside a physician’s primary specialty.^26^ Furthermore, GPT models show promise in identifying rare diseases that might otherwise be overlooked and in recommending appropriate referrals to specialized care.^27^ In medically underserved areas or environments with limited access to specialist consultation, AI systems can help reduce healthcare disparities by providing consistent decision support and expanding diagnostic reach.^28^

### Remaining challenges and ethical considerations

Although AI demonstrates high diagnostic accuracy, the role of clinicians remains indispensable. The selection, preprocessing, and input of relevant patient information into AI systems depend heavily on clinician expertise.^29^ Moreover, while the models have demonstrated impressive performance based on the provided case information, real clinical settings are far more complex.^30^ Patients often provide incomplete histories or omit critical information, presenting substantial challenges that current LLMs are not yet fully equipped to manage.^31^ Further prospective studies using real-world clinical data, including free-form questions and open-ended scenarios, are recommended to evaluate the robustness and generalizability of these models.

As AI technologies, including large multimodal models capable of analyzing imaging data, become increasingly integrated into clinical workflows, new ethical considerations must be addressed. In addition to traditional concerns about accountability, transparency, and bias in algorithmic decision-making, the inclusion of sensitive medical images heightens the importance of patient privacy and data protection.^32^ Furthermore, establishing clear legal frameworks is essential to delineate responsibility in cases where AI-assisted decisions result in misdiagnosis.^33^ Despite the substantial capabilities demonstrated by these models, it remains crucial to recognize that AI systems currently lack the emotional understanding, empathy, and holistic grasp of individual patient cases that clinicians possess; thus, human intervention and interaction remain essential.

### Limitations of the study

Our study has certain limitations. First, clinicians were defined as site voters rather than as a standardized group of certified specialists, and the voting participants included a broad range of users. The absence of participant credentials and the informal nature of responses, provided without academic or clinical consequences, may have contributed to an underestimation of actual clinician performance. Despite these limitations, the primary aim of the study was not to benchmark against definitive human performance but to illustrate the rapid and consistent progress of LLMs in performing medical tasks.

Given the observed performance trajectory suggesting increasing clinical applicability, further studies involving board-certified clinicians are warranted to evaluate the models’ validity in real-world settings. Second, the dataset was derived from Medscape clinical case challenges, which may not fully capture the diversity and complexity of real-world scenarios. Additionally, because the clinical case dataset is open-source, it is possible that the GPT models were exposed to specific cases during training. However, o1 and GPT-4o were trained only on data available until October 2023, and their accuracy rates showed no statistically significant differences between datasets introduced before and after this date, suggesting that any prior exposure had a limited impact on model accuracy.

Third, the dataset may have been influenced by inherent bias. This includes selection bias, as Medscape’s educational objectives often prioritize complex or rare conditions, leading to their preferential selection over common clinical presentations. Furthermore, latent biases, such as the overrepresentation of specific medical conditions, demographic groups, or clinical scenarios, may have limited the dataset’s generalizability and reduced its ability to reflect real-world patient diversity.^34^

## CONCLUSIONS

The o1 and GPT-4o models achieved higher accuracy than clinicians on clinical case challenge questions, including those involving medical imaging. These findings suggest that both models can serve as valuable tools for healthcare professionals, supporting various aspects of patient care and decision-making in clinical settings.

## Data Availability

All data produced are available online at https://reference.medscape.com/features/casechallenges

## RESOURCE AVAILABILITY

### Lead contact

Further information and requests for resources and reagents should be directed to and will be fulfilled by the lead contact, SungA Bae.

### Materials availability

This study did not generate new materials.

### Data and code availability

The publicly available Medscape case challenge questions used for this analysis are available for download at the cited sources: https://reference.medscape.com/features/casechallenges. The underlying code and sample datasets used in this study are publicly available in the following GitHub repository: https://github.com/makogirls/GPT-medscape-case-challenge.

## ACKNOWLEDGMENTS

This research was supported by a grant from the Korea Health Technology R&D Project through the Korea Health Industry Development Institute (KHIDI), funded by the Ministry of Health and Welfare, Republic of Korea (grant number RS-2023-KH135442).

Additionally, this study was supported by a New Faculty Research Seed Money Grant from Yonsei University College of Medicine for 2025 (2025-32-0022).

## AUTHOR CONTRIBUTIONS

Conceptualization, S. Bae and J.Y. Park; funding acquisition, S. Bae and J.Y. Park; supervision, S. Bae and J.Y. Park; data curation, J. Jung and H. Kim; formal analysis, J. Jung and H. Kim; investigation, J. Jung and H. Kim; programming, J. Jung and H. Kim; verification, J. Jung and H. Kim; visualization, J. Jung and H. Kim; writing—original draft, all authors; writing—review & editing, all authors; final approval, all authors; accountability, all authors.

## DECLARATION OF INTERESTS

None.

## DECLARATION OF GENERATIVE AI AND AI-ASSISTED TECHNOLOGIES

During the preparation of this manuscript, GPT-4o and o1 (OpenAI, 2025) were used as supplementary tools to assist with translation, refine sentence structure, and generate vector images for figures. These tools were not involved in generating original scientific content, nor did they contribute to the interpretation of the study results.

## SUPPLEMENTAL INFORMATION

**Document S1. Figures S1–S2 and Tables S1–S2**

**Figure S1. Medscape quiz sample**

**Figure S2. Model Performances Before and After the Knowledge Cutoff Date**

**Table S1. Comparison of clinician and GPT model responses**

**Table S2. Medscape quiz information**

## STAR⍰ METHODS

### KEY RESOURCES TABLE

| REAGENT OR RESOURCE | SOURCE | IDENTIFIER |
| --- | --- | --- |
| Deposited data |  |  |
| Medscape case challenge questions | https://reference.medscape.com/features/casechallenges | The Medscape case challenge scenarios and questions are publicly available on the website. |
| Software and algorithms |  |  |
| Python | https://www.python.org/ | Version 3.11.3 |
| pandas | https://pandas.pydata.org/ | Version 2.2.2 |
| openai | https://platform.openai.com/docs | Version 1.86.0 |
| GPT-3.5 Turbo | https://platform.openai.com/docs/models/gpt-3-5 | Version gpt-3.5-turbo-0613 |
| GPT-4 Turbo | https://platform.openai.com/docs/models/gpt-4 | Version gpt-4-0613 |
| GPT-4 Omni | https://platform.openai.com/docs/models/gpt-4o | Version gpt-4o-2024-05-13 |
| o1 | https://platform.openai.com/docs/models/o1 | Default Version (queried in Dec 2024) |
| R | https://www.r-project.org/ | Version 4.5.0 |

## EXPERIMENTAL MODEL AND STUDY PARTICIPANT DETAILS

### Study Design

This retrospective study analyzed Medscape clinical case challenge questions from May 2011 to June 2024 (**Figure 4** and **Table S2 in the Supplementary Appendix**).^35^ Medscape clinical case challenges are interactive educational tools designed to help healthcare professionals assess and improve their clinical knowledge and decision-making skills. Exclusion criteria included non-question contents and multiple choice questions. Case data were collected using a custom Python-based web scraper that retrieved publicly available content from the Medscape website. Only publicly accessible cases were included, and no personal data were collected. Ethical approval was not required due to the retrospective nature of the study and the use of open-source data.

**Figure 4.**
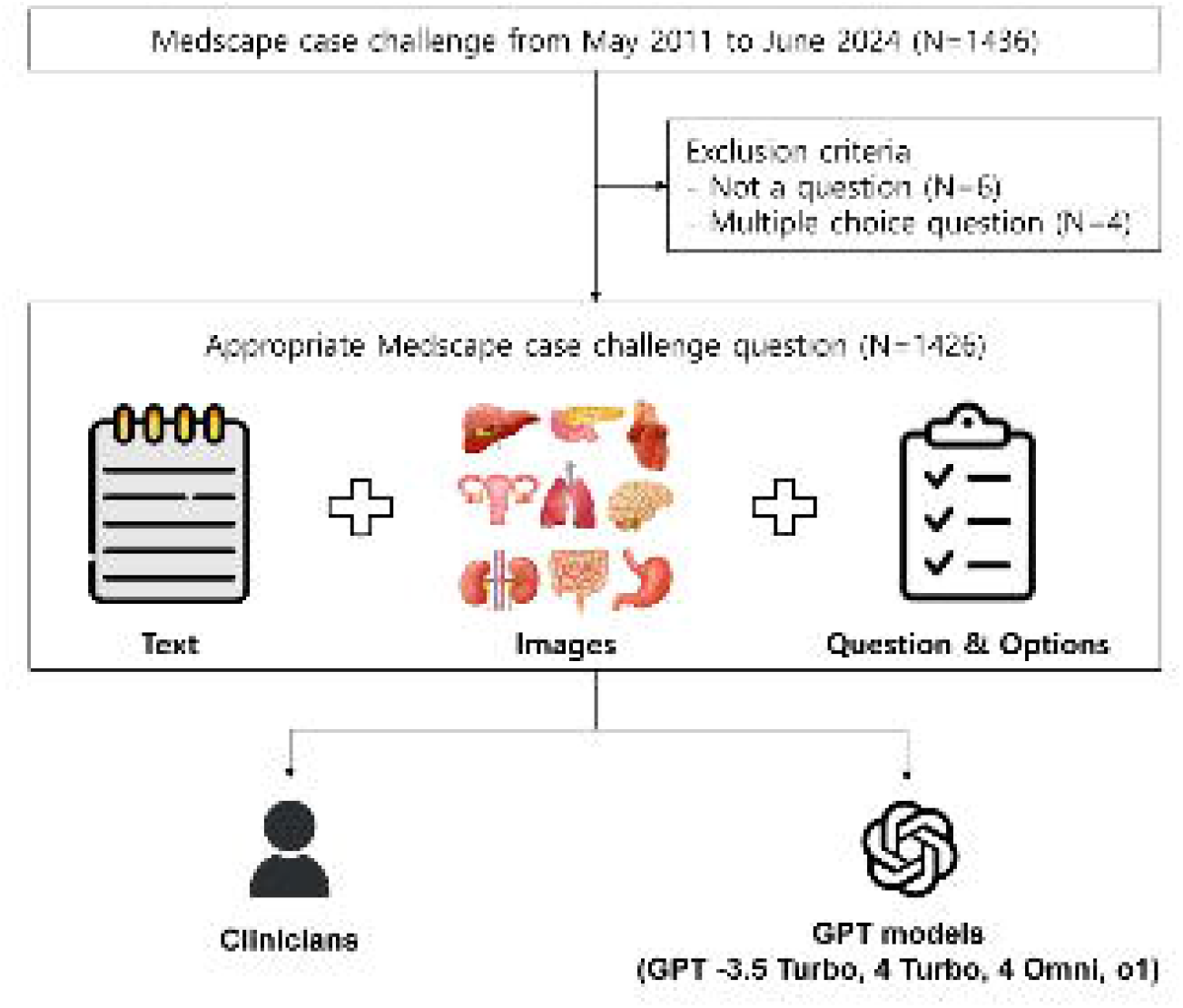
Study flowchart. This flowchart illustrates the workflow for evaluating the diagnostic performances of GPT models compared with clinicians in Medscape case challenge questions. GPT, Generative Pre-trained Transformer

Each case presented a realistic clinical scenario, including patient history, physical examination findings, diagnostic test results, and imaging studies. Participants were tasked with selecting the most appropriate answer from a set of multiple-choice options covering aspects of diagnosis, disease characteristics, examinations, and treatment strategies. Clinicians’ responses were defined as the most frequently selected option for each question. Each question had a predefined correct answer established by the Medscape case author based on the confirmed diagnosis and clinical guidelines. The cases were categorized by medical specialty, including internal medicine (allergy, cardiology, endocrinology, gastroenterology, hematology, infectious diseases, nephrology, oncology, pulmonology, and rheumatology), major surgery (obstetrics/gynecology and surgery), pediatrics, psychiatry, and minor specialties (dermatology, emergency medicine, neurology, neurosurgery, ophthalmology, orthopedics, otorhinolaryngology, and urology).

## METHOD DETAILS

### Large Language Models and Prompt

Each GPT model (GPT-3.5 Turbo, GPT-4 Turbo, GPT-4 Omni, and o1) was accessed through the OpenAI application programming interface (API) between August and December 2024. Eash model was tasked to generate a formatted response based on the options available for each question. To ensure consistency and replicability, the models were queried using predefined prompt templates that simulated the clinical reasoning scenarios. **Table 3** outlines the prompts used to query the models, including their structures and formatting. For questions that included images, both textual and visual data were provided to the models, where applicable, enabling multimodal input processing in GPT-4o and o1. The original images uploaded by Medscape were used without any preprocessing. The chat session was reset before each new question to eliminate the influence of memory retention or in-context learning, ensuring that all questions were processed independently and that responses remained unbiased. GPT-3.5 Turbo, GPT-4 Turbo, and GPT-4 Omni were configured with a temperature of 0.1 and top-p of 1.0, while o1 used its default settings of a temperature of 1.0 and top-p of 1.0.

**Table 3.** Prompt template for GPT models

| Models | Prompt |
| --- | --- |
| GPT-3.5 Turbo<br>GPT-4 Turbo | You are a specialist clinician. The data provided is about one patient, and it might involve a rare disease, so consider many possibilities. As a clinician, you must make data-based decisions, highly recommended to be based on journals and medical textbooks. Examination findings are very important!<br>[Question]<br>[Options]<br>[All textual information] |
| GPT-4 Omni<br>o1 | You are a specialist clinician. The data provided is about one patient, and it might involve a rare disease, so consider many possibilities. As a clinician, you must make data-based decisions, highly recommended to be based on journals and medical textbooks. Examination findings are very important!<br>[Question]<br>[Options]<br>[All textual information]<br>[Images] |

## QUANTIFICATION AND STATISTICAL ANALYSIS

We compared the performances of the four GPT models (GPT-3.5 Turbo, GPT-4 Turbo, GPT-4 Omni, and o1) with those of clinicians using mixed-effects logistic regression analysis to evaluate differences in accuracy. The model type was included as a fixed effect to compare performance across models, and a random intercept was added for each case to account for potential correlations among nested questions within the same case. We calculated the accuracy and 95% confidence intervals (CIs) for each model and clinician and performed pairwise comparisons between them. For cases with images, a focused analysis of GPT-4o and o1 was conducted to specifically assess their multimodal capability in interpreting visual data. The dataset was further analyzed by question category (diagnosis, characteristics, examination, and treatment) and medical specialty (internal medicine, major surgery, pediatrics, psychiatry, and minor specialties). GPT-4o and o1 were tested over five independent trials using the same set of questions to evaluate response consistency. All analyses were conducted using R (version 4.5.0, R Foundation for Statistical Computing, Vienna, Austria), with a *P*-value < .05 considered statistically significant.

## Notes

### Competing Interest Statement

The authors have declared no competing interest.

### Author Declarations

This study used only publicly available data.

